# Excess Out-Of-Hospital Mortality and Declining Oxygen Saturation: The Sentinel Role of EMS Data in the COVID-19 Crisis in Tijuana, Mexico

**DOI:** 10.1101/2020.05.13.20098186

**Authors:** Joseph Friedman, Alhelí Calderón-Villarreal, Ietza Bojorquez, Carlos Vera Hernández, David L. Schriger, Eva Tovar Hirashima

## Abstract

**Objective:** Emergency medical services (EMS) may serve as a key source of real-time data about the evolving health of COVID-19 affected populations, especially in low-and-middle-income countries (LMICs) with less rapid and reliable vital statistic registration systems. Although official COVID-19 statistics in Mexico report almost exclusively in-hospital mortality events, excess out-of-hospital mortality has been identified in other settings, including one EMS study in Italy that showed a 58% increase. EMS and hospital reports from several countries have suggested that silent hypoxemia—low oxygen saturation (SpO2) in the absence of dyspnea—is associated with COVID-19 outbreaks. It is unclear, however, how these phenomena can be generalized to LMICs. We assess how EMS data can be used in a sentinel capacity in Tijuana, a city on the Mexico-United States border with earlier exposure to COVID-19 than many LMIC settings.

**Methods:** We calculated numbers of weekly out-of-hospital deaths and respiratory cases seen by EMS in Tijuana, and estimate the difference between peak-epidemic rates (during April 14^th^-May 11^th^) and forecasted 2014–2019 trends. Results were compared with official COVID-19 statistics, stratified by neighborhood socioeconomic status (SES), and examined for changing demographic or clinical features, including mean (SpO2).

**Results:** An estimated 194.7 (95%CI: 135.5-253.9) excess out-of-hospital deaths events occurred, representing an increase of 145% (70%-338%) compared to forecasted trends. During the same window, only 8 COVID-19-positive, out-of-hospital deaths were reported in official statistics. This corresponded with a rise in respiratory cases of 274% (119%-1142%), and a drop in mean SpO2 to 77.7%, from 90.2% at baseline. The highest out-of-hospital death rates were observed in low-SES areas, although respiratory cases were more concentrated in high-SES areas.

**Conclusions:** EMS systems may play an important sentinel role in monitoring excess out-of-hospital mortality and other trends during the COVID-19 crisis in LMICs. Using EMS data, we observed increases in out-of-hospital deaths in Tijuana that were nearly threefold greater magnitude than increases reported using EMS data in Italy. Increased testing in out-of-hospital settings may be required to determine if excess mortality is being driven by COVID-19 infection, health system saturation, or patient avoidance of healthcare. We also found evidence of worsening rates of hypoxemia among respiratory patients seen by EMS, suggesting a rise in silent hypoxemia, which should be met with increased detection and clinical management efforts. Finally, we observed that social disparities in out-of-hospital death that warrant monitoring and amelioration.

## Introduction

As the coronavirus disease, 2019 (COVID-19) spreads across most countries of the world, real-time information is required to detect and manage the health of the population. This is a particular challenge in low-and-middle-income countries (LMICs), such as Mexico, due to less rapid and robust vital statistic registration systems. Although a vital registration system does exist in Mexico, official statistics are available on an approximately 2-year lag, and records of mortality are not always reliable—a condition similar to most LMICs^1-3^. As of May 2020, no official mortality statistics for Mexico were available beyond the year 2018^4^. Given these data restrictions, information from emergency medical services (EMS) may serve as a key source of real-time knowledge about the evolving health of COVID-19 affected populations, offering information of clinical significance.

EMS data can play a particular role in measuring out-of-hospital mortality. As the epidemiological properties of the COVID-19 pandemic have become more clear, excess mortality has become an important area of study. A small number of analyses have been published—initially largely by news organizations—describing excess total mortality^5-7^. However, due to the aforementioned limitations, no data from Mexico, or the vast majority of LMICs, were available as of May 2020. Out-of-hospital deaths represent an important facet of total excess mortality, which may be particularly suited for measurement using EMS data. One recent report from the Lombardy region of Italy used EMS records to show an increase of 58% compared to prior year values, during the peak of the epidemic^8^. It is unclear, however, how this phenomenon would play out in LMICs with relatively weaker health systems^9-13^. In the context of COVID-19, an increase in out-of-hospital mortality could be expected, directly from COVID-19, or indirectly as patients delay care and health systems become overwhelmed^14-16^. Nevertheless, rates of out-of-hospital mortality remain a generally understudied facet of the pandemic^6,7,17^, and to our knowledge, there is little evidence for LMICs.

Another key area that can be monitored using data from EMS systems during the COVID-19 pandemic is the detection of “silent hypoxia”. Reports from China, and later Italy, the US, and Norway, have described many COVID-19 patients who initially present with hypoxemia without signs of respiratory distress (“silent hypoxemia”) and later go on to develop respiratory failure^18-21^. It is possible that this kind of hypoxemia, and subsequent rapid decompensation^22^, could result in mortality before patients are able to access EMS or hospital services, especially in areas were health systems are saturated or patients are not able to quickly access healthcare services when decompensation occurs.

Mexico is a middle-income country that saw its first confirmed case of COVID-19 on February 27^th^, and reached 10,000 cases by April 17^th^, according to official statistics^23^. Tijuana, in Northern Mexico, is a city of over 1.7 million inhabitants that shares a heavily crossed border with San Diego County, California, in the United States^24^. As of May 12^th^, the international border remained open to residents of the United States, although Mexican nationals with tourist visas were generally barred from crossing beginning in late March. Tijuana therefore may have been subjected to earlier exposure to SARS-CoV-2 than the rest of Mexico due to the importation of cases from California^25,26^. Reported cases of COVID-19 in Tijuana were among the first in Mexico—beginning on March 17^th^. On May 11th Tijuana had the highest number of COVID-19 deaths of any municipality in the country (170), and the mortality rate (17.3 per 100,000 people) was almost six times the national rate of 3.1 per 100,000 people^23,24,27^. Therefore, Tijuana may represent an important bellwether for the rest of Mexico and have general relevance to trends that will be experienced by the EMS systems of other LMICs.

Using EMS data from Tijuana, our primary objective was to describe the potential sentinel role for EMS data in monitoring the epidemiological profile of the COVID-19 epidemic in a LMIC context. We focused the analysis on trends in out-of-hospital mortality and silent hypoxemia among respiratory patients. We also sought to characterize any changes in demographics, geography, and neighborhood socioeconomic status (SES) among these patient groups. Additionally, we aimed to compare trends documented by the EMS system with official government statistics describing COVID-19 cases and deaths.

## Methods

### Study design and setting

We used data from the Mexican Red Cross in Tijuana, which responds to approximately 98% of 9-1-1-activations of EMS care in the city^28^. We drew upon routinely collected, deidentified, encounter-level records describing patient characteristics and the provision of emergency medical services. We conducted a retrospective, descriptive analysis comparing the observed peak epidemic to prior trends. We excluded calls that were cancelled before the ambulance arrived at the scene. Given that rates of violence in Tijuana have been highly variable in recent years, complicating the estimation of expected trends, we also excluded patients suffering from traumatic injuries from all analyses. Data were available for most of the January 2014 through May 11^th^, 2020, although some records, including files from 2018 and February 2020, were not available in digital form on the rapid timescale required to conduct this analysis. Publicly available data describing official confirmed cases and deaths stemming from COVID-19 were obtained from the Mexican National Office of Epidemiology^29^. This study was deemed exempt from review by the University of California, Los Angeles Institutional Review Board.

### Measures

Out-of-hospital mortality was defined as a case in which a patient was found dead-on-arrival, or died before reaching a hospital, as documented by EMS. We also assessed the number of cases of respiratory morbidity. This was defined as either a) a chief complaint of “respiratory”, “difficulty breathing”, or “respiratory infection” (which collectively represented the majority of cases) or b) a chief complaint that was metabolic or gastrointestinal in nature, combined with an SpO2 of less than 92%. The decision to include gastrointestinal or metabolic patients with low SpO2 reflected recent reports of atypical COVID-19 presentations with chiefly gastrointestinal symptoms^30^ as well as the association with diabetes mellitus^31^. In all cases, if a series of SpO2 measurements were taken, we used the first available value, taken before treatment began.

For cases of out-of-hospital mortality, we assessed patient age, gender, health insurance status (including uninsured, privately insured, or membership in one of several main public healthcare systems), time from call to ambulance arrival, if CPR was administered, neighborhood of residence, and administrative geostatistical-area level SES. For respiratory cases, we assessed the aforementioned variables as well as level of consciousness and SpO2.

The neighborhood *(colonia)* of residence was mapped using a shapefile from the Mexican National Population Council (CONAPO). An index of SES *(índice de marginación)* and populations were provided at the level of basic geostatistical area (AGEB, in Spanish) defined by the Mexican Institute of Statistics and Geography (INEGI)^32,33^, which typically include several neighborhoods, and are based on 2010 census data. We created a categorical SES variable, defined as population-weighted quintiles of the continuous SES variable, categorized as “lowest”, “low”, “medium”, “high”, and “highest”. As neighborhoods and AGEB do not overlap perfectly, a linkage was performed between neighborhood and AGEB, in order to assign SES values to each neighborhood. This involved finding the midpoint of each neighborhood and assigning it the SES value of the basic statistical unit where it was located. In the small number of cases where the midpoint of a neighborhood fell outside of a defined AGEB, the neighborhood cluster was assigned to the closest AGEB to the midpoint.

Official data describing COVID-19 cases and deaths in Tijuana were aggregated to weekly totals, and graphed alongside EMS-documented numbers.

### Analysis

Changes in out-of-hospital mortality was assessed by comparing weekly statistics from January through May of 2020, to forecasted values estimated using baseline trends from January 1st, 2014 to December 31st, 2019. The process was repeated for the primary outcome measures (number of out-of-hospital deaths, number of respiratory cases) as well as two outcomes assessed as sensitivity analyses (proportion of cases that result in out-of-hospital mortality, proportion of cases that are respiratory in nature) to control for potential differences in case volume. Using OLS regression we modelled the seasonal time trend with a fixed effect dummy variable on each week of the year. The secular trend was captured using a linear continuous fixed effect on year. Forecasts with 95% prediction intervals were made by extrapolating the model through May 2020. Ratios of observed to expected numbers and proportions, and their uncertainty intervals were calculated by divided the observed value in each week by the forecasted value and prediction interval. We compared pre-epidemic SpO2 values with those seen during the peak epidemic period. We also described trends in the distribution of SpO2 during the epidemic, as measured by quintiles of the distribution of SpO2, and examined the relationship between SpO2 and level of consciousness.

We also sought to ensure that no difference in nomenclature, classification, or life support practices occurred in response to the onset of the COVID-19 crisis that could cause an apparent increase in out-of-hospital mortality. We therefore assessed rates of cardiopulmonary resuscitation (CPR), ambulance transit times, and the total composition of all cases, before and during the COVID-19 period.

All cases were included in the sections of the analysis for which they had available data. Missing values are noted as applicable in Tables 1 and 2.

## Results

### Overall Profile of EMS Cases

The total number of cases was relatively similar during the peak observed COVID period, with 410 average weekly cases between April 14^th^ and May 11^th^, compared to a weekly mean of 382.9 in 2019 (Figure 1). We noted a dropping quantity of non-urgent cases, which fell to 39.0% of all cases April 14^th^ and May 11^th^, as compared to a 59.1% average for 2019, likely due to social distancing and increased reluctance to use healthcare services for non-urgent matters. Contrastingly, both urgent and deceased cases rose, reaching 11.2% and 20.0% respectively, as compared to 6.7% and 7.9% respectively in 2019.

**Figure 1.**
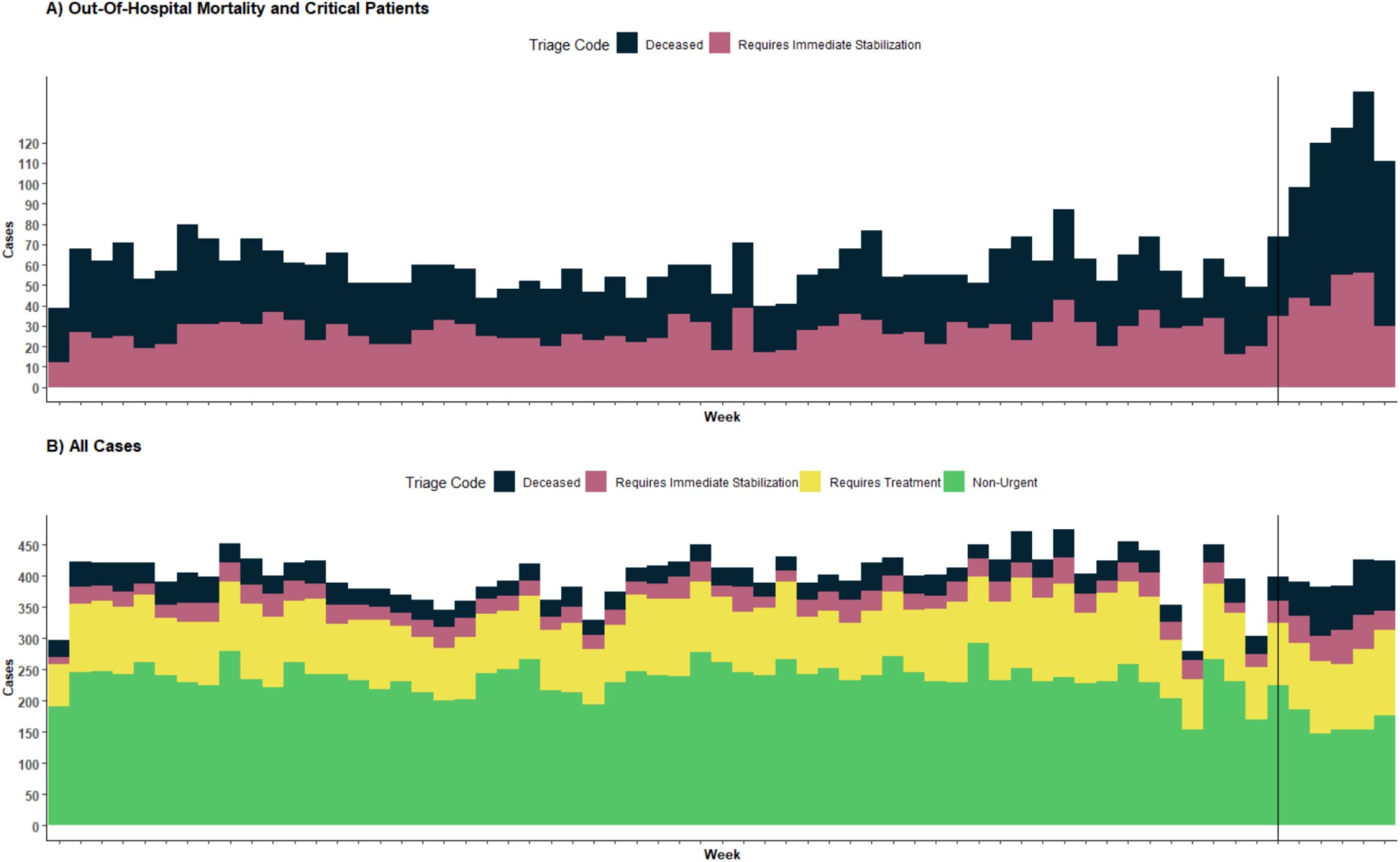
Weekly Case Breakdown by Triage Priority Code, 2019-2020. Part A shows only pre-hospital mortality cases and patients in critical condition who require urgent hospitalization. Part B shows the full distribution patients. Both parts refer to non-trauma patients, and include data from 2019 through April 2020. The vertical black line marks the week of March 31^st^, when respiratory morbidity cases began to rise.

There were no substantial differences in CPR rates before or during the COVID-19 period (Table 1), likely because overall CPR administration rates were generally quite low among non-trauma patients in Tijuana (Supplemental Figure 1). Average ambulance travel time from call to arrival-on-scene was slightly longer during the observed COVID-19 peak period (20.5 minutes) compared to 2019 (16.4 minutes). Of note, pre-epidemic transit times were higher than those typically seen in higher-income urban areas, and may help explain low life-support rates among critical patients.

### Out-Of-Hospital Mortality

From January to March 2020, the number and proportion of out-of-hospital mortality cases was within or below the 95% prediction interval based on trends observed from 2014 to 2019 (Figure 2). However, the week of April 14^th^ 80 deaths (Figure 3, Part A) exceeding the previously observed maximum in the timeseries (Figure 2). The 329 deaths occurring from April 14^th^ to May 11^th^ were compared to the predicted number of 134.7 (95%CI: 75.1-193.5) for the same period, yielding an estimated excess of 194.7 (95%CI: 135.5-253.9) deaths. This represents an increase of 145% (70%-338%) compared to forecasted trends. Similar results were seen when modelling the percent of cases represented by deaths, and restricting the analysis to only dead-on-arrival pre-hospital mortality (see Supplemental Figures 3 and 2, respectively). The peak observed period of out-of-hospital mortality lined up exactly with the highest observed rates of COVID-19 deaths according to official statistics (Figure 3, Part C). 262 deaths among confirmed COVID-19 patients were reported during the same period. However, only 8 of these deaths were reported as occurring “in an outpatient context”, the remainder being reported as occurring among “hospitalized patients”^29^.

**Figure 2.**
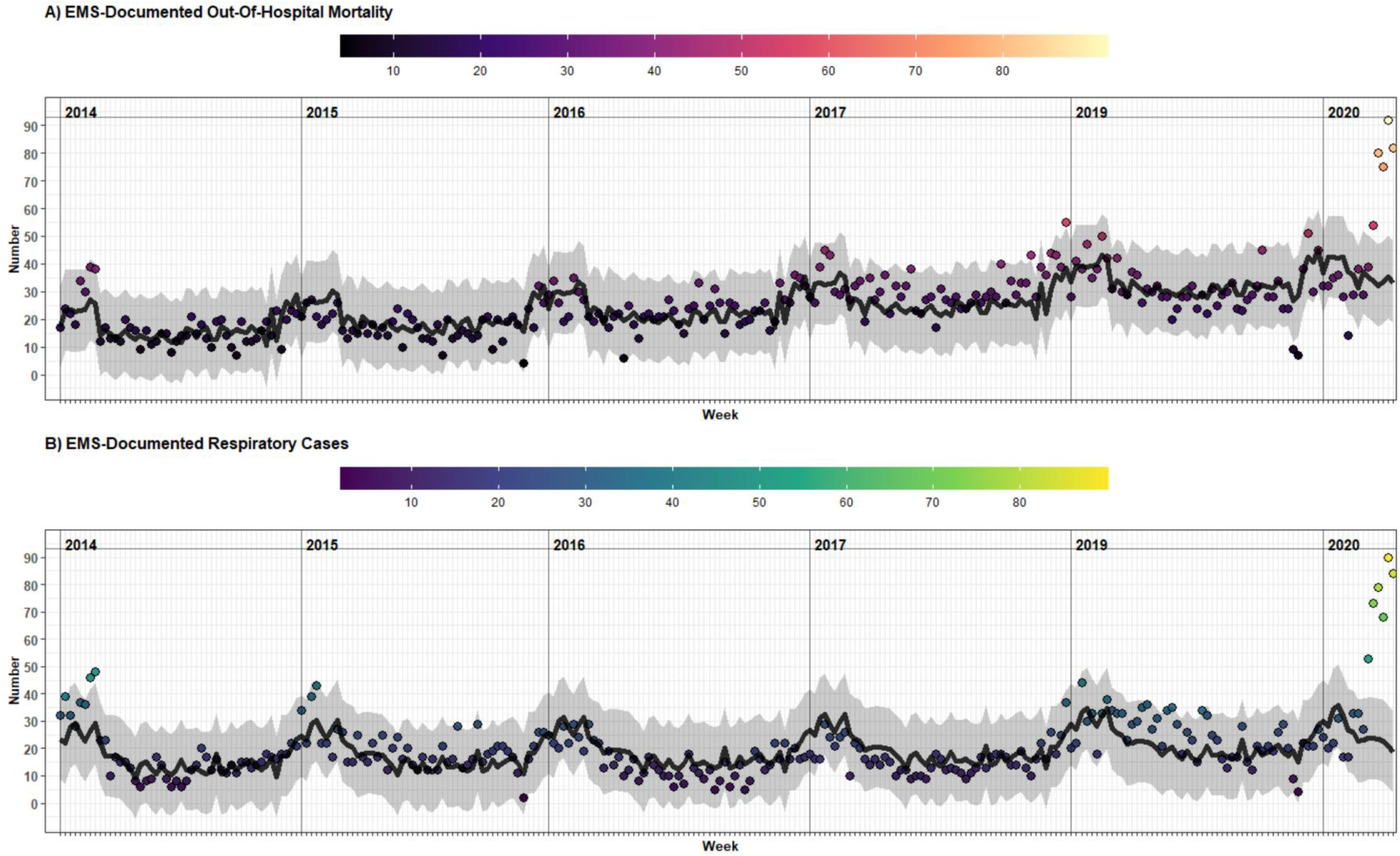
Long-Run EMS-Documented Out-Of-Hospital Mortality and Respiratory Cases, 2014-2020. A) EMS-documented out-of-hospital mortality. B) EMS-documented respiratory cases. Parts A and B include expected values (black line) and 95% prediction intervals (grey band) based on model fit on data from 2014-2019, with forecasts through May of 2020. Both series exclude trauma-patients.

**Figure 3.**
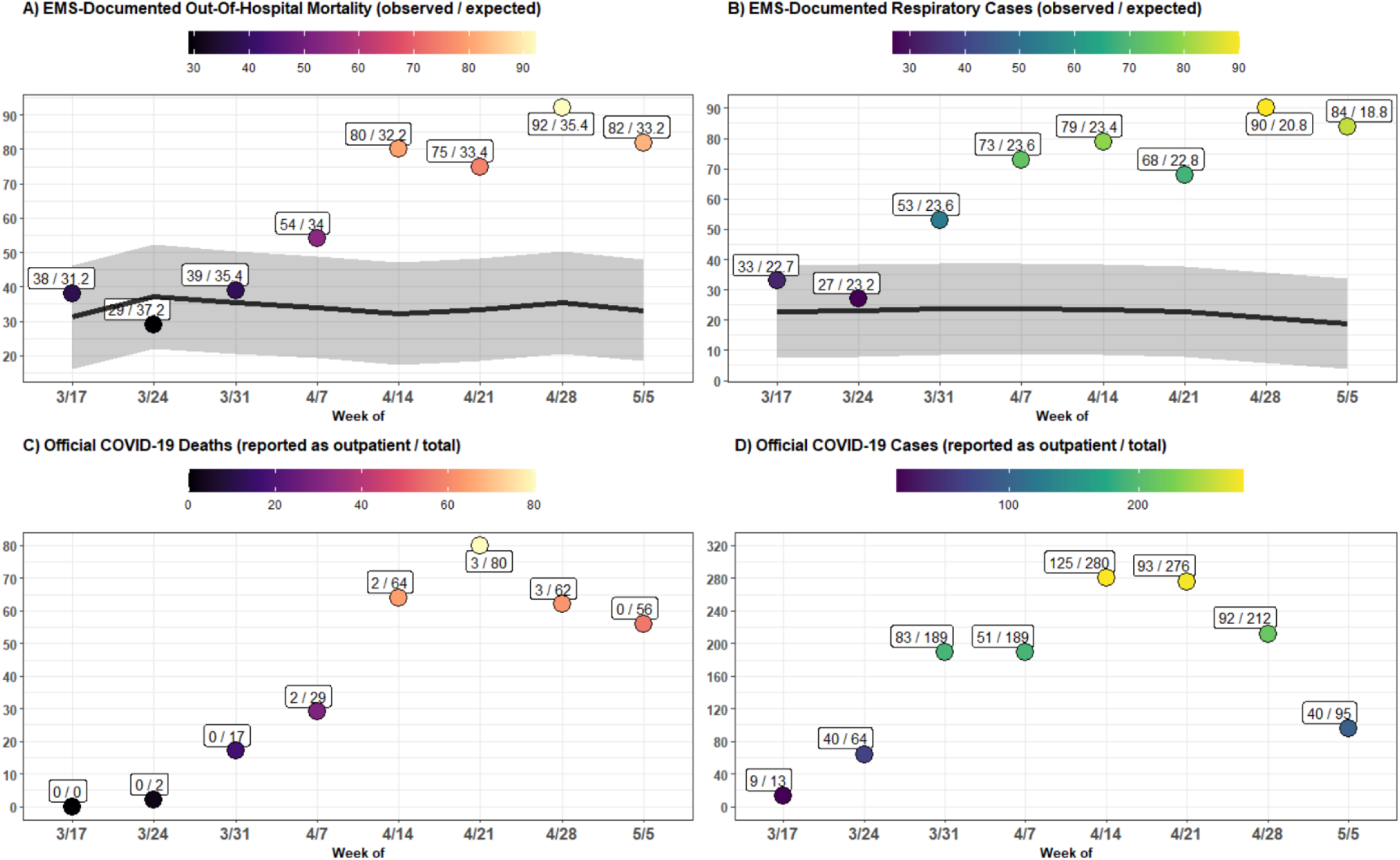
EMS-Documented Out-Of-Hospital Mortality and Respiratory Cases Compared to Official COVID-19 Case and Mortality Numbers, March 17^th^ - May 11^th^. A) EMS-documented out-of-hospital mortality, with the observed and expected number shown in text. B) EMS-documented respiratory cases, with the observed and expected number shown in text. C) Deaths among patients with confirmed COVID-19, according to official national government statistics, with the number reported as managed in the outpatient setting and the total reported in text. D) Number of patients with confirmed COVID-19, according to official national government statistics, with the number reported as managed in the outpatient setting and the total reported in text. Parts A-D refer to weekly totals. Parts A and B include expected values (black line) and 95% prediction intervals (grey band) based on forecasted trends from 2014-2019. Parts A and B exclude trauma patients.

Out-of-hospital deaths during the period of April 14^th^ to May 11^th^ were majority men (68.4%), of working age 18-64 (64.1%), who were beneficiaries of the Mexican National Institute for Social Security (IMSS) healthcare system (45.9%) (Table 1). IMSS is a social security scheme providing health care to individuals employed in the private formal sector. Although the age and gender patterns were largely similar to those observed throughout 2019, they were more likely to be IMSS beneficiaries, (45.9% vs. 29.9%, difference=16.3% [95%CI: 10.0%-22.0%]).

**Table 1.** Characteristics of Out-Of-Hospital Mortality Patients.

|  | All 2019<br>(N=1569) | April 14<sup>th</sup> to May 11th<br>(N=329) |
| --- | --- | --- |
| **Age** |  |  |
| Mean (SD) | 59.2 (20.1) | 58.0 (17.4) |
| Median [Min, Max] | 60.0 [1.00, 104] | 59.0 [1.00, 97.0] |
| Missing | 3 (0.2%) | 0 (0%) |
| **Categorical Age** |  |  |
| 0-18: Pediatric | 43 (2.7%) | 5 (1.5%) |
| 18-64: Adult | 871 (55.5%) | 211 (64.1%) |
| 65+: Senior | 652 (41.6%) | 113 (34.3%) |
| Missing | 3 (0.2%) | 0 (0%) |
| **Gender** |  |  |
| Female | 499 (31.8%) | 104 (31.6%) |
| Male | 1070 (68.2%) | 225 (68.4%) |
| **Health Insurance** |  |  |
| IMSS | 469 (29.9%) | 151 (45.9%) |
| ISSSTE* | 51 (3.3%) | 6 (1.8%) |
| Uninsured | 686 (43.7%) | 140 (42.6%) |
| Seguro Popular/INSABI** | 285 (18.2%) | 20 (6.1%) |
| Private Insurance | 78 (5.0%) | 12 (3.6%) |
| **CPR** |  |  |
| Advanced CPR | 21 (1.3%) | 1 (0.3%) |
| Basic CPR | 47 (3.0%) | 4 (1.2%) |
| None | 1501 (95.7%) | 324 (98.5%) |
| **Call-to-Arrival Time** |  |  |
| Mean (SD) | 16.4 (10.1) | 20.5 (12.6) |
| Median [Min, Max] | 15.0 [0, 190] | 19.0 [0, 135] |
| Missing | 65 (4.1%) | 26 (7.9%) |

Numbers exclude trauma-related deaths. The peak observed mortality period, from April 14<sup>th</sup> to May 11th, is compared to all of 2019.
\*Health insurance for government workers (*Instituto de Seguridad y de Servicios Sociales de los Trabajadores del Estado*)
\*\*Social safety net health insurance (*Instituto Nacional de Salud para el Bienestar*)

### Respiratory Morbidity and Oxygen Saturation

In addition to out-of-hospital mortality, we also noted an increase in respiratory cases, which rose above the prediction interval of expected values during the week of March 31^st^ (Figure 3, part B). Nevertheless, the peak observed to expected ratio occurred in the same week as that of out-of-hospital mortality, reaching 90 cases on the week of April 28^th^. During the April 14^th^ to May 11^th^ window, 321 respiratory cases were observed, representing 235.1 (175.1-295.1) more than expected, an increase of 274% (120%-1141%). Similar results were seen when modelling the percent of respiratory cases (see Supplement Figure 3). Similar to the trends observed for out-of-hospital mortality, respiratory patients during the March 31^st^ to May 11^th^ period were majority men (61.5%), of working age (72.0%), and IMSS beneficiaries (66.4%) (Table 2). Compared with respiratory patients in 2019, respiratory patients in the peak observed epidemic period more likely to be IMSS beneficiaries (66.4% vs. 38.9%, difference=27.6% [22.3%-32.9%]), and have an SpO2 lower than 90% (54.8% vs. 32.4%, difference=22.4% [17.0%-27.9%]).

**Table 2.** Characteristics of Respiratory Patients by Week.

|  | March 31<sup>st</sup><br>(N=53) | April 7th<br>(N=73) | April 14th<br>(N=79) | April 21st<br>(N=67) | April 28th<br>(N=90) | May 5<sup>th</sup><br>(N=84) | All 2019<br>(N=1253) |
| --- | --- | --- | --- | --- | --- | --- | --- |
| **Age** |  |  |  |  |  |  |  |
| Mean (SD) | 55.8 (18.6) | 51.3 (17.1) | 49.6 (18.9) | 53.8 (15.6) | 55.1 (15.9) | 57.8 (14.5) | 53.5 (24.6) |
| Median [Min, Max] | 55.5 [1.00, 95.0] | 51.0 [19.0, 89.0] | 44.0 [1.00, 94.0] | 52.5 [2.00, 97.0] | 56.0 [4.00, 88.0] | 58.0 [28.0, 92.0] | 56.0 [1.00, 105] |
| Missing | 1 (1.9%) | 0 (0%) | 0 (0%) | 0 (0%) | 0 (0%) | 0 (0%) | 2 (0.2%) |
| **Categorical Age** |  |  |  |  |  |  |  |
| 0-18: Pediatric | 1 (1.9%) | 0 (0%) | 1 (1.3%) | 1 (1.5%) | 2 (2.2%) | 0 (0%) | 118 (9.4%) |
| 18-64: Adult | 37 (69.8%) | 59 (80.8%) | 57 (72.2%) | 51 (75.0%) | 63 (70.0%) | 55 (65.5%) | 666 (53.2%) |
| 65+: Adult Mayor | 14 (26.4%) | 14 (19.2%) | 21 (26.6%) | 16 (23.5%) | 25 (27.8%) | 29 (34.5%) | 467 (37.3%) |
| Missing | 1 (1.9%) | 0 (0%) | 0 (0%) | 0 (0%) | 0 (0%) | 0 (0%) | 2 (0.2%) |
| **Gender** |  |  |  |  |  |  |  |
| Female | 24 (45.3%) | 30 (41.1%) | 24 (30.4%) | 28 (41.2%) | 29 (32.2%) | 37 (44.0%) | 569 (45.4%) |
| Male | 29 (54.7%) | 43 (58.9%) | 55 (69.6%) | 40 (58.8%) | 61 (67.8%) | 47 (56.0%) | 684 (54.6%) |
| **Health Insurance** |  |  |  |  |  |  |  |
| IMSS | 35 (66.0%) | 54 (74.0%) | 56 (70.9%) | 42 (61.8%) | 57 (63.3%) | 53 (63.1%) | 487 (38.9%) |
| ISSSTE* | 2 (3.8%) | 1 (1.4%) | 0 (0%) | 3 (4.4%) | 2 (2.2%) | 6 (7.1%) | 46 (3.7%) |
| Uninsured | 11 (20.8%) | 11 (15.1%) | 16 (20.3%) | 18 (26.5%) | 23 (25.6%) | 18 (21.4%) | 310 (24.7%) |
| Seg.Pop / INSABI** | 2 (3.8%) | 4 (5.5%) | 4 (5.1%) | 5 (7.4%) | 5 (5.6%) | 5 (6.0%) | 359 (28.7%) |
| Private | 3 (5.7%) | 3 (4.1%) | 3 (3.8%) | 0 (0%) | 3 (3.3%) | 2 (2.4%) | 51 (4.1%) |
| **CPR** |  |  |  |  |  |  |  |
| Advanced CPR | 0 (0%) | 0 (0%) | 0 (0%) | 0 (0%) | 0 (0%) | 0 (0%) | 3 (0.2%) |
| Basic CPR | 0 (0%) | 1 (1.4%) | 1 (1.3%) | 1 (1.5%) | 0 (0%) | 0 (0%) | 7 (0.6%) |
| None | 53 (100%) | 72 (98.6%) | 78 (98.7%) | 67 (98.5%) | 90 (100%) | 84 (100%) | 1243 (99.2%) |
| **Call-to-Arrival Time** |  |  |  |  |  |  |  |
| Mean (SD) | 18.1 (10.4) | 28.7 (63.3) | 19.8 (9.62) | 21.3 (9.05) | 19.8 (14.7) | 22.4 (13.4) | 16.4 (26.1) |
| Median [Min, Max] | 17.0 [0, 61.0] | 17.0 [0, 517] | 18.5 [0, 42.0] | 20.0 [8.00, 43.0] | 18.0 [0, 88.0] | 21.5 [0, 67.0] | 14.0 [0, 620] |
| Missing | 0 (0%) | 8 (11.0%) | 11 (13.9%) | 10 (14.7%) | 13 (14.4%) | 4 (4.8%) | 57 (4.5%) |
| **Level of Concious.** |  |  |  |  |  |  |  |
| Alert | 46 (86.8%) | 60 (82.2%) | 67 (84.8%) | 48 (70.6%) | 72 (80.0%) | 64 (76.2%) | 930 (74.2%) |
| Verbal Stimulus | 2 (3.8%) | 3 (4.1%) | 2 (2.5%) | 4 (5.9%) | 8 (8.9%) | 2 (2.4%) | 65 (5.2%) |
| Painful Stimulus | 2 (3.8%) | 4 (5.5%) | 3 (3.8%) | 5 (7.4%) | 3 (3.3%) | 9 (10.7%) | 107 (8.5%) |
| Unresponsive | 0 (0%) | 1 (1.4%) | 3 (3.8%) | 3 (4.4%) | 2 (2.2%) | 0 (0%) | 78 (6.2%) |
| Missing | 3 (5.7%) | 5 (6.8%) | 4 (5.1%) | 8 (11.8%) | 5 (5.6%) | 9 (10.7%) | 73 (5.8%) |
| **SpO2** |  |  |  |  |  |  |  |
| Mean (SD) | 89.0 (10.4) | 83.1 (15.4) | 83.1 (14.6) | 81.8 (16.2) | 77.1 (19.8) | 77.7 (18.5) | 90.0 (10.6) |
| Median [Min, Max] | 90.0 [50.0, 99.0] | 88.0 [40.0, 98.0] | 86.5 [30.0, 100] | 88.0 [43.0, 100] | 82.0 [0, 98.0] | 83.0 [31.0, 98.0] | 92.0 [0, 100] |
| Missing | 3 (5.7%) | 4 (5.5%) | 3 (3.8%) | 6 (8.8%) | 5 (5.6%) | 6 (7.1%) | 65 (5.2%) |

Numbers exclude trauma-related patients and pre-hospital deaths. The peak observed respiratory case period, from March 31<sup>st</sup> to May 11th, is compared to all of 2019.
\*Health insurance for government workers (*Instituto de Seguridad y de Servicios Sociales de los Trabajadores del Estado*)
\*\*Social safety net health insurance (*Instituto Nacional de Salud para el Bienestar*)

The overall trend of EMS-documented respiratory cases was quite similar to that observed in confirmed COVID-19 cases reported in official statistics (Figure 3, Part D). However, the magnitude was substantially lower; the week of April 14^th^, for example, saw 280 COVID-19 cases, 155 of which were hospitalized, which greatly exceeded the 79 total EMS-documented respiratory cases. Additionally, a falling number of confirmed COVID-19 cases was observed in the April 28^th^ to May 11^th^ period, and no corresponding drop was seen among EMS-observed respiratory cases.

The mean SpO2 value among respiratory patients declined steadily, from 90.0% during the pre-epidemic period of 2019, reaching a low of 77.7% during the week of April 28^th^ (Table 2). Figure 4 shows the weekly evolution of the distribution of SpO2 values among respiratory patients. The highest quintile of the distribution of SpO2 values remained fairly stable throughout the study period, with a median value above 95%. Nevertheless, the remaining quintiles of the distribution generally decreased in their SpO2 values, and a widening of the distribution of SpO2 was observed as a result. In the week of April 7^th^, the lowest quintile of the distribution of SpO2 values fell sharply, reaching a median SpO2 of 55%. Notably, despite the lower average SpO2, the proportion of patients presenting as alert and oriented did not see a commensurate drop relative to baseline (table 2), even among the lowest quintile of SpO2 values (Figure 4).

**Figure 4.**
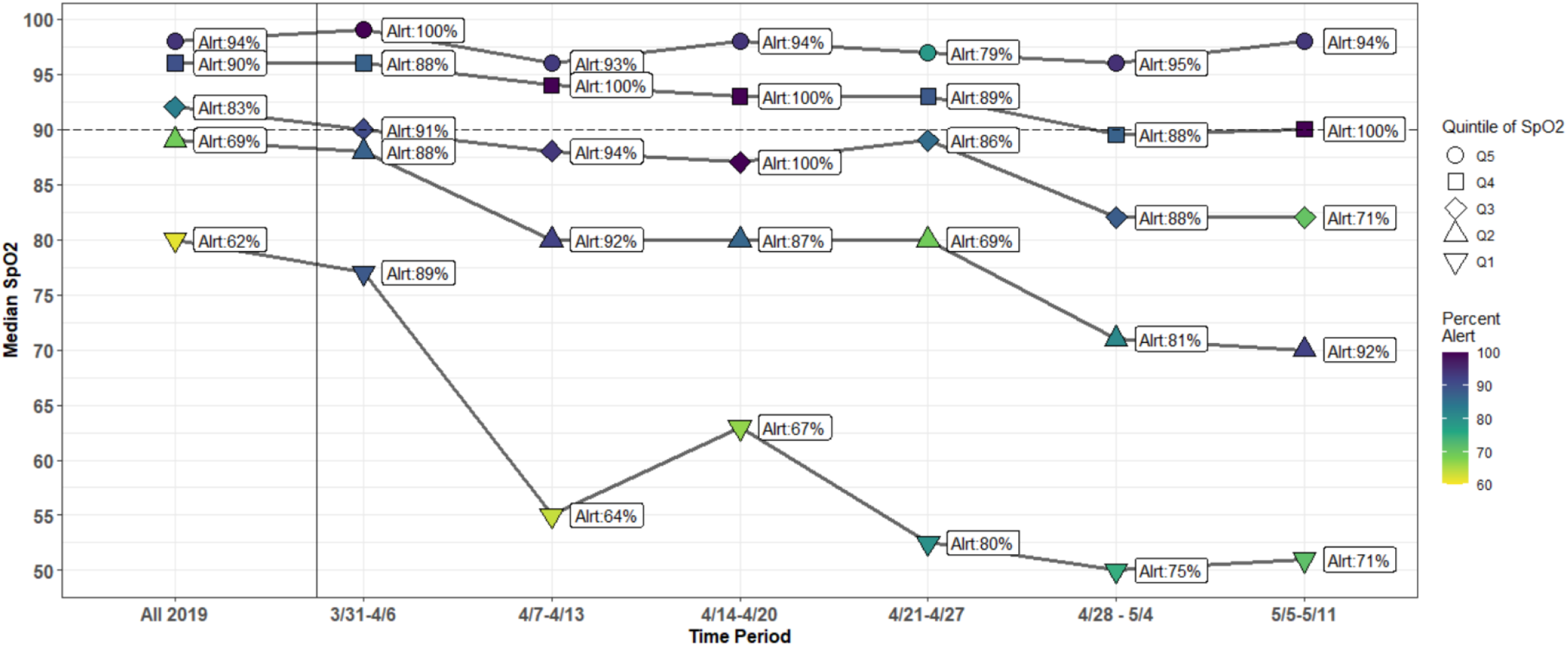
Trends in SpO2 and Percent Presenting Alert Among EMS-Documented Respiratory Cases. The distribution of SpO2 values over time is visualized weekly from March 31^st^ to May 11th, 2020 and compared to all data from 2019. Respiratory cases were divided into 5 quintiles of SpO2 values, and the median of each quartile is plotted. The color reflects the percent of individuals in each quartile that presented as alert, which is also plotted as text next to each point.

### Socioeconomic Status

EMS data can also provide insights into the location of outbreaks and to social disparities in the distribution of the COVID-19 mortality and morbidity Figure 5 highlights the SES and geospatial distribution of out-of-hospital mortality and respiratory cases in Tijuana. It is noteworthy that the largest clusters of out-of-hospital mortality did not occur in the same locations as the largest clusters of respiratory cases. We observe that clusters of respiratory cases during the peak epidemic period were most concentrated in highest-and high-SES quintiles of Tijuana. Contrastingly, the largest clusters of out-of-hospital mortality cases were seen in the low-SES quintile. As rates per 100,000 people the low-SES quintile of the population saw the highest rate of out-of-hospital mortality at 24.5, while the high-SES quintile saw the highest rate of respiratory cases, at 30.9.

**Figure 5.**
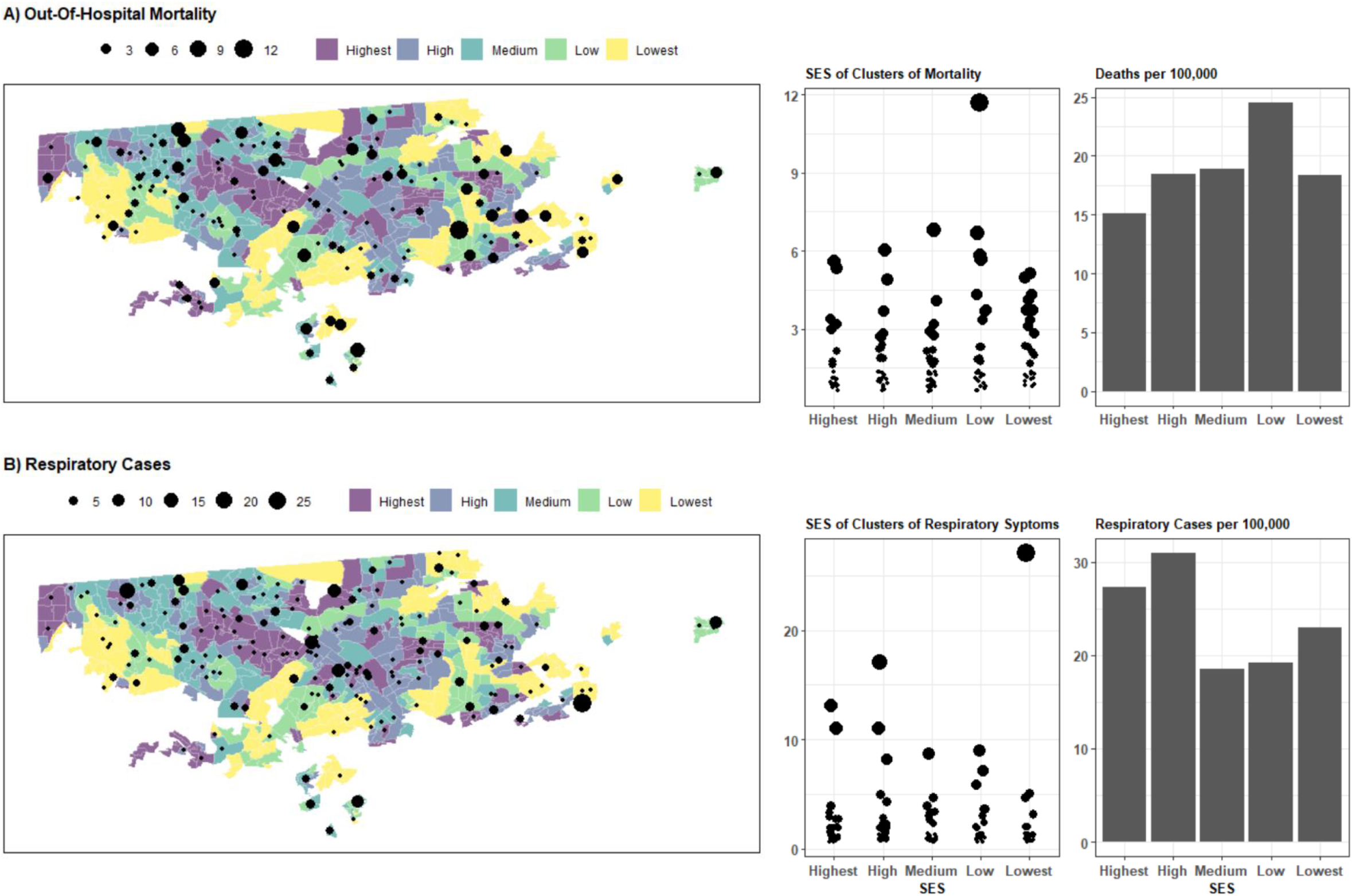
Out-Of-Hospital Mortality and Respiratory Cases by Neighborhood and Neighborhood SES. The categorical socioeconomic status (SES) of each basic statistical unit *(ageb)* is mapped for Tijuana. Overlaid is the out-of-hospital mortality occurring during April 14^th^ to May 11^th^ (part A) and respiratory cases occurring during March 31^st^ to May 11^th^ (part B). The number of cases in each neighborhood *(colonia)* is shown as a point, with the size reflecting the magnitude. In the middle column, the points are organized by neighborhood SES. On the right, the number of cases is shown as a rate per 100,000 people, for each quintile of neighborhood SES.

### Limitations

We are unable to differentiate whether the excess out-of-hospital mortality is solely attributable to COVID-19 infections, or if it also reflects increased death rates from other causes. For example, increased cardiac arrest frequency could arise if patients stayed home during ischemic chest pain episodes, because the system was saturated, or they were afraid to seek care. Although we did examine the diagnostic codes associated with each death, the EMTs completing the records were unable to reliably ascertain the cause of death in the majority of cases, and information about prior COVID-19 tests was typically not available. Similarly, our measure of respiratory cases only reflects SpO2 values below 92%, or other respiratory symptoms, and cannot directly indicate patient COVID-19 status. Like any analysis using EMS data, the out-of-hospital mortality statistics presented here cannot capture events occurring in the absence of 9-1-1 activations. Though we note a large increase in out-of-hospital mortality, our results may not represent the full in increase in absolute numbers. However, if the proportion of deaths resulting in 9-1-1 activations were to be correlated with the onset of the COVID-19 pandemic, that could bias our results in an unpredictable direction.

We use neighborhood-level SES which is an imperfect measure of person-level SES. Furthermore, we use population and SES values from the most recent census, 2010, as newer data were not available. This may miss trends in some rapidly changing parts of Tijuana. This analysis should be updated when 2020 census data become available. Additionally, the model we used to extrapolate past trends into 2020 was straightforward in design, and we did not perform out-of-sample predictive validity testing or compare alternative predictive model forms. Nevertheless, given the magnitude of the disparities observed, and the presence of some missing data in past years of observed trends, we opted for a simple and easy-to-interpret model. Furthermore, we note that many studies of this nature simply use the prior year’s values as a comparison group^8^, and therefore a simple approach may be preferable.

### Discussion

We used data from Tijuana, Mexico to illustrate how EMS systems may be a useful source of real-time information for tracking the COVID-19 epidemic in LMICs contexts where other sources of information are not rapidly available. We showed that out-of-hospital mortality documented by the EMS system increased dramatically during the peak observed COVID-19 epidemic period seen in April and May 2020. The relative excess mortality—145% above baseline—represents between a two and-threefold higher magnitude increase compared to the 58% figure reported in a recent similar study from the Lombardy region of Italy^8^. This may be related to Tijuana being in a middle-income country, with a relatively more fragile healthcare system and lower-income population. These results suggest that other regions of Mexico, and LMICs in general, may need to plan for and ameliorate sharply increasing rates of out-of-hospital mortality in order to prevent a large burden of potentially unmeasured death stemming from the COVID-19 pandemic. These findings echo a growing number of calls for health system strengthening in LMIC in the face of the COVID-19 pandemic^9-13^.

During the April 14^th^ to May 11^th^ period in which we estimate 194.7 excess deaths occurred, only 8 official COVID-19 deaths were reported as “outpatient”, the remainder being categorized as “hospitalized”. This suggests that the increase in out-of-hospital mortality that we observed cannot be explained by official COVID-19 statistics. Importantly, we were not able to ascertain the etiology of the excess mortality we observe. It is therefore possible that most of the excess deaths resulted from non-COVID-19 causes of death, stemming from delay of care or health system saturation. It is also possible that many of the deaths we observe represent COVID-19 patients who were never diagnosed or formally tracked as such. Finally, delays in reporting or data presentation may simply have led to lower weekly totals for out-of-hospital mortality among known COVID-19 patients who are not hospitalized. In any case, EMS data represent an important source of near-real-time information that can be used to rapidly track the evolving health of COVID-19 affected populations. We propose that EMS systems may play an especially important sentinel role in LMICs, and can be used to monitor excess out-of-hospital mortality during the COVID-19 crisis. This function may be of particular importance in LMICs given the lack of access to rapid vital registration records.

Notably, in the most recent two weeks of official statistics, a drop in confirmed COVID-19 cases and deaths was observed. However, the number of EMS-documented out-of-hospital deaths and total respiratory cases did not show a corresponding decrease. More research is required to explore what role access to COVID-19 tests, lags in official COVID-19 statistics, or access to hospital beds, may be playing in driving differences between EMS-documented, and official statistics. Increased testing in out-of-hospital settings may be required to determine if excess mortality is being driven by COVID-19 infection, health system saturation, or patient avoidance of healthcare.

Although EMS staff were not able to generate substantial clinical information about patients who died before reaching a hospital, as most were found dead-on-arrival, important clues about the etiology of out-of-hospital mortality can be gleaned by assessing clinical and demographic characteristics among living patients seen for respiratory symptoms during the same period. During the window of observed peak excess mortality, we also observed a concurrent elevation in the rate of patients presenting with respiratory symptoms. These patients had similar demographic characteristics to the out-of-hospital mortality cases. Although the number of respiratory patients reached the highest rate observed during the 2014-2020 period studied, they were still lower than the number of official COVID-19 patients that were hospitalized. This suggests that most officially documented COVID-19 patients are reaching healthcare facilities independently of the EMS system in Tijuana.

The detection of silent hypoxemia is difficult by definition. Patients typically present to EMS services only after they experience dyspnea. Nevertheless, it is possible that some indirect evidence about silent hypoxemia can be observed in the declining SpO2 values seen among respiratory patients during the observed peak COVID-19 window. We saw a sharp decline in mean SpO2 values, although no concomitant decrease was seen among the percentage of patients who were alert on presentation. Hypoxemia is a known predictor of mortality among COVID-19 patients^34^, and these data may suggest that silent hypoxemia and subsequent rapid decomposition is a relevant factor to understanding out-of-hospital mortality rates.

Given the novel nature of the COVID-19 pathophysiology, more education about silent hypoxemia is needed for physicians to better manage it clinically, and patients to better understand the risks^22,34^. As COVID-19 quickly overwhelms frail health-systems, clinicians on the frontlines may easily overlook a “well appearing” patient despite a low SpO2, to make room for patients who are overtly sick. It is important for patients to understand that in silent hypoxemia, dyspnea is a late-stage symptom, and their condition may be deteriorating without perceived decreases in subjective respiratory ability^19^. Detection of hypoxemia in the general population should be undertaken, and priority areas can be identified using clusters from EMS data, such as those shown in Figure 5 of this text. For resource-limited settings, a potential screening tool can be found in the ROTH scale proposed by Chorin et al.^35^ It is a simple breathing and counting exercise that has been shown to have a positive correlation with measured SpO2. It was originally conceived as a tool for telemedicine, and has not been validated in patients with COVID. However, in resource-limited settings, it may also be administered in-person in lieu of a pulse-oximeter or via phone. To facilitate it’s calculation, a free open access website application was created in Spanish on April 30^th^, 2020 (https://pruebaroth.com/)^36^. The scale is imperfect and it will require validation among COVID-19 patients to be used widely; however, it may provide some triage benefit in settings where wide-scale measurement using pulse oximetry is not feasible.

The social pattern of out-of-hospital mortality and respiratory cases also deserves consideration and monitoring in the COVID-19 crisis context. We observed a differential trend by neighborhood-SES between out-of-hospital mortality and respiratory cases. Although respiratory cases were strikingly concentrated in the high- and highest-SES quintiles, the highest out-of-hospital mortality rates were observed in low-SES areas. There is a notable difference between respiratory cases and deaths, which may suggest that the profile of individuals who have the economic or social capital to seek care early for respiratory symptoms in Tijuana differs from those who do not interact with the medical system until after their death. This finding adds to a growing body of literature and social commentary suggesting that social inequalities may be translating into inequalities in the risk of infection or death from COVID-19 in numerous contexts^37-43^

### Conclusions

EMS data provide a valuable tool to rapidly track the health of populations at risk of COVID-19 in LMICs, where other forms of real-time data may not be available. EMS information can be used to track excess out-of-hospital mortality and respiratory burden, as well as changing clinical or demographic features.

Detected clusters of out-of-hospital mortality or cases can be subsequently targeted for screenings for hypoxemia and/or COVID-19 status. Social disparities in COVID-19 and out-of-hospital mortality should be monitored, and additional resources may need to be directed to low-SES areas within LMIC cities.

Among respiratory patients, the drop of SpO2 observed during the peak epidemic period suggests that hypoxemia precedes clinical manifestations such as dyspnea, a term coined “silent hypoxemia”. The lack of overt clinical manifestations early in the disease, and the resulting difficulty of detecting silent hypoxemia, may be a driver of out-of-hospital mortality in LMICs where the health system is easily overwhelmed, and accessing EMS services is more difficult.

## Data Availability

Data describing confirmed COVID-19 cases and Deaths are publicly available at: https://www.gob.mx/salud/documentos/datos-abiertos-152127. EMS data may be requested from the Mexican Red Cross by contacting the study corresponding author [ETH].

## Author Contributions

JF, ACV and ETH conceived and designed the study, and oversaw data acquisition. JF and ACV analyzed the data. All authors contributed to data interpretation as well as drafting and critical revision of the manuscript.

## Acknowledgements

The authors thank Alberto Luna for his efforts related to data preparation and management. We thank the Mexican Red Cross, Dr. Andres Smith, Juan Carlos Mendez, and the numerous EMTs and paramedics working tirelessly on the front lines, for facilitating data collection and access.

